# Decoding resistance: interpretable machine learning to predict ciprofloxacin resistance in *Shigella* spp

**DOI:** 10.64898/2026.04.07.26350353

**Authors:** Mahmood R. Gohari, Pauline Zhang, Andre Villegas, Laura C. Rosella, Samir N. Patel, Jessica P. Hopkins, Venkata R. Duvvuri

## Abstract

Antimicrobial resistance (AMR) is a growing global public health threat that complicates the treatment and control of bacterial infections. *Shigella* spp., a leading cause of bacterial diarrhea worldwide, has increasingly exhibited resistance to multiple antimicrobial agents that are commonly recommended therapy for severe shigellosis. Although conventional antimicrobial susceptibility testing (AST) remains the reference standard, it is time-consuming and provides limited insight into the genetic mechanisms underlying resistance. Whole-genome sequencing (WGS) has emerged as a complementary approach for AMR detection by enabling direct identification of resistance genetic determinants encoded in bacterial genomes. Machine learning (ML) methods applied to genomic features such as k-mers have shown promise for predicting resistance phenotypes from WGS data; however, applications to *Shigella* remain limited. In this study, we developed and evaluated an interpretable ML framework for predicting ciprofloxacin resistance using k-mer features derived from WGS data of 1,424 *Shigella* isolates collected in Ontario, Canada, between 2018 and 2025. K-mers were extracted from known gene targets associated with ciprofloxacin resistance, including chromosomal quinoline resistance-determining regions (QRDRs: *gyrA* and *parC*) and plasmid-mediated determinants (qnr).

Supervised ML approaches were trained and compared. We evaluated the influence of k-mer lengths (k=11, 15, 21 and 31) on predictive performance and model interpretability; and compared models based on chromosomal determinants alone and models incorporating both chromosomal and plasmid-mediated determinants. Randon Forest classifier achieved the most consistent performance across models. Inclusion of plasmid-mediated determinants improved predictive accuracy relative to chromosomal-only models. Although differences across k-mer lengths were modest, k = 11 produced the highest area under the receiver operating characteristic curve (AUC) and the lowest Brier score. SHAP analyses localized high-impact features within QRDRs of *gyrA* and *parC,* supporting biological interpretability. These findings demonstrate that biologically-informed k-mer–based ML models can accurately and transparently predict ciprofloxacin resistance in *Shigella,* supporting their potential integration into genomic AMR surveillance and digital public health frameworks.

**Author summary:** In this study, we used genome sequencing data to develop machine learning models that predict ciprofloxacin resistance for *Shigella* directly from bacterial DNA. We focused on small DNA fragments (k-mers) derived from known resistance genes and mutations. Among the approaches tested, a Random Forest model showed the most consistent performance. Combining chromosomal mutations with plasmid-mediated resistance genes improved prediction accuracy and helped identify key genetic regions associated with resistance. These findings demonstrate that machine learning applied to genomic data can accurately and interpretable predict antibiotic resistance, supporting its potential use in genomic surveillance and public health monitoring.

## Background

Antimicrobial resistance (AMR) is a major and growing global public health threat, contributing to an estimated 4.9 million deaths annually (1). The increasing prevalence of resistance limits effective treatment options, prolongs illness, facilitates transmission of infectious diseases, and increases healthcare costs (2, 3). *Shigella* spp., a leading cause of bacterial gastroenteritis worldwide and responsible for substantial morbidity, particularly among children and other vulnerable populations (4), has emerged as a pathogen of increasing concern in the context of rising AMR (1). Rising resistance in *Shigella* complicates clinical management and poses growing challenges for public health surveillance. Resistance to multiple first-line agents, including ampicillin, azithromycin, and ciprofloxacin, has increasingly been reported, further limiting treatment options for Shigellosis (5). Reflecting the growing global threat of AMR, the World Health Organization (WHO) has identified *Shigella* as a priority pathogen requiring enhanced surveillance and the development of new antimicrobial strategies (6). The increasing prevalence of resistant *Shigella* strains therefore represents a significant therapeutic and public health challenge, highlighting the need for rapid and reliable approaches to monitor and predict resistance patterns.

In current practice, AMR detection relies primarily on conventional phenotypic antimicrobial susceptibility testing (AST), which remains the reference standard but is time-consuming and resource-intensive, often delaying optimal therapy (7, 8). In addition, phenotypic AST provides limited insight into genetic mechanisms underlying resistance and is not well suited for the early detection of emerging or uncommon resistance patterns (8). In addition to AST, whole-genome sequencing (WGS) has emerged as a complementary approach, enabling direct interrogation of the known genetic determinants of resistance. By capturing both resistance genes and point mutations associated with antimicrobial agents, WGS provides a more comprehensive characterization of AMR mechanisms and supports genomic surveillance of bacterial pathogens (9, 10). Because AMR commonly arises through mechanisms such as point mutations, gene amplification, and horizontal gene transfer, WGS can detect multiple resistance determinants simultaneously from a single assay (10). Given the growing availability and significance of WGS data for AMR surveillance, machine learning (ML) approaches have increasingly been applied to predict resistance phenotypes from genomic data, with studies reporting high concordance (>95%) between genotypic predictions and AST results for major bacteria (11, 12). ML models have been applied to genomic features derived from WGS data, such as single nucleotide polymorphisms (SNPs) and k-mers, to predict AMR. These high-dimensional genomic representations are particularly well suited to ML approaches because they enable the identification of complex and potentially non-linear relationships between genomic features and resistance phenotypes(13, 14). These capabilities have led to increasing interest in ML-based genomic prediction as a complementary approach for multiple bacterial pathogens, including *Escherichia coli*, *Klebsiella pneumoniae*, and *Staphylococcus aureus*(*11, 15–17*), However, their application to *Shigella* spp. remain limited.

Ciprofloxacin is a commonly recommended empiric treatment for severe shigellosis; however, resistance has increasingly been reported in *Shigella* (5). Ciprofloxacin resistance in *Shigella* is driven by chromosomal mutations, in the quinolone resistance–determining regions (QRDR) of *gyrA* and *parC* genes, which encode DNA gyrase and topoisomerase IV, as well as plasmid-mediated resistance genes such as *qnr* (18, 19). K-mer–based genomic features are widely used in ML models for AMR prediction, but an important methodological consideration is the choice of k-mer length, as it can influence both predictive performance and biological resolution (20).

Previous studies have suggested that optimal k-mer length is context-dependent and may vary across datasets, pathogens, and analytical objectives (20). In this study, we developed and evaluated a ML framework to predict ciprofloxacin resistance in *Shigella* using k-mer–based genomic features derived from WGS data of isolates collected in Ontario, Canada, between 2018-2025. The primary objective was to compare the performance of established ML models for predicting ciprofloxacin resistance. We further examined how k-mer length influences predictive performance and model interpretability; and compared models based on chromosomal determinants alone (*gyr*A, *par*C) with those incorporating both chromosomal and plasmid-mediated determinants (*qnr*).

## Results

Our dataset comprised *Shigella* spp. isolates representing species circulating in Ontario, predominantly *S. sonnei* (51.7%, n=743) and *S. flexneri* (45.9%, n=659), with smaller proportions of *S. boydii* (0.8%, n=13) and *S. dysenteriae* (0.6%, n=9).

### Distribution and Co-occurrence of Ciprofloxacin Variants

The distribution of ciprofloxacin antimicrobial susceptibility phenotypes by combinations of known ciprofloxacin genetic resistance determinants (gyrA, parC, and qnr) is presented in Table 1. Each variant (v1 to v6) represents a unique combination based on the presence (+) or absence (−) of these genetic resistance determinants. Isolates with mutations in the chromosomal genes gyrA and parC, with or without the presence of qnr, accounted for the majority of phenotypically resistant isolates (89.9%; n = 515/577). In contrast, isolates lacking mutations in both chromosomal genes were phenotypically susceptible (99.8%, n =659/660) (Table 1). The presence of *qnr* in the absence of mutations in the chromosomal genes *gyr*A and *par*C was predominantly associated with susceptible phenotypes. Among the 292 isolates carrying *qnr* without mutations in *gyrA* and *parC*, 119 (40.8%) were phenotypically susceptible, 166 (56.8%) intermediate, and only 7 (2.4%) as resistant.

**Table 1.**
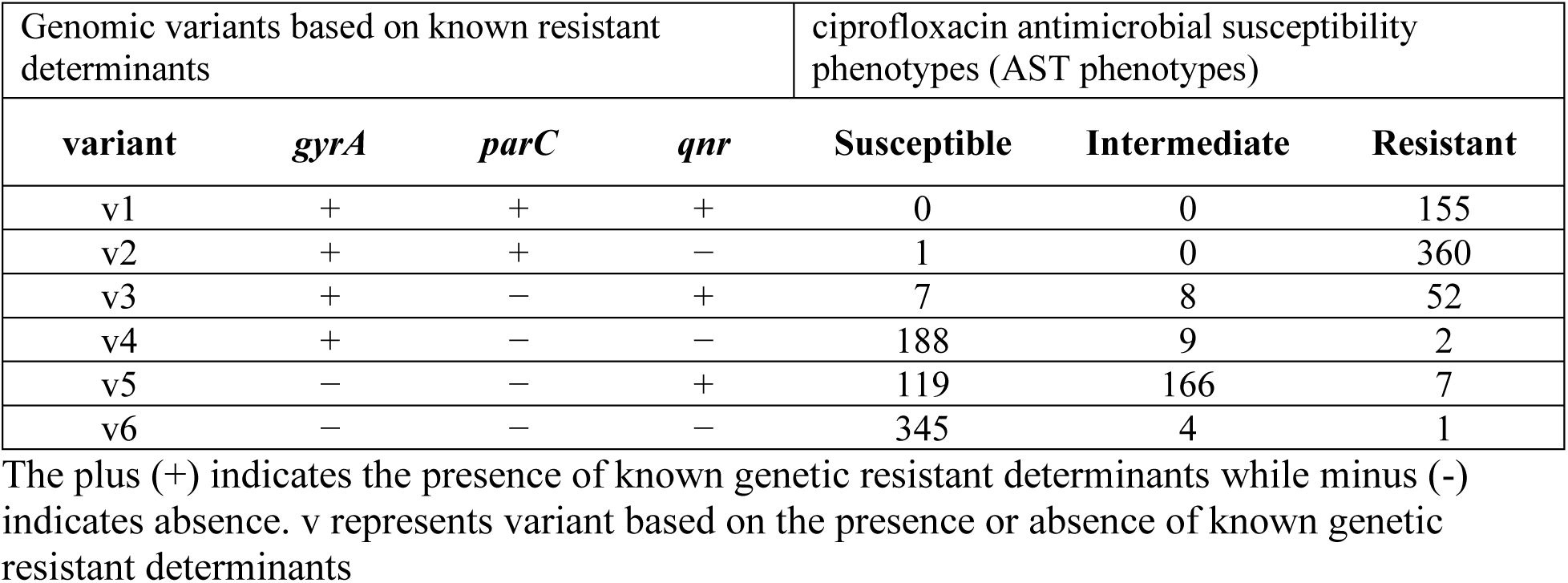
Distribution of ciprofloxacin antimicrobial susceptibility phenotypes by combinations of known ciprofloxacin genetic resistant determinants (*gyr*A, *par*C, and *qnr*) among clinical isolates collected in Ontario between 2018 and 2025.

Figure 2 illustrates the co-occurrence patterns among key ciprofloxacin resistance determinants across ciprofloxacin known gene variants. Strong correlations were observed among canonical QRDR mutations in *gyrA* and *parC*, particularly between *gyrA* S83 and D87 amino acid (AA) substitutions and *parC* S80I AA substitution, which clustered predominantly within the variant (*gyrA+, parC+, qnr+*). In contrast, susceptible isolates (*gyrA*-, *parC-, qnr*-) were largely characterized by wild-type or single-mutation profiles. Plasmid-mediated *qnr* subtypes exhibited weak correlations with chromosomal QRDR mutations, suggesting largely independent acquisition pathways.

**Figure 1.**
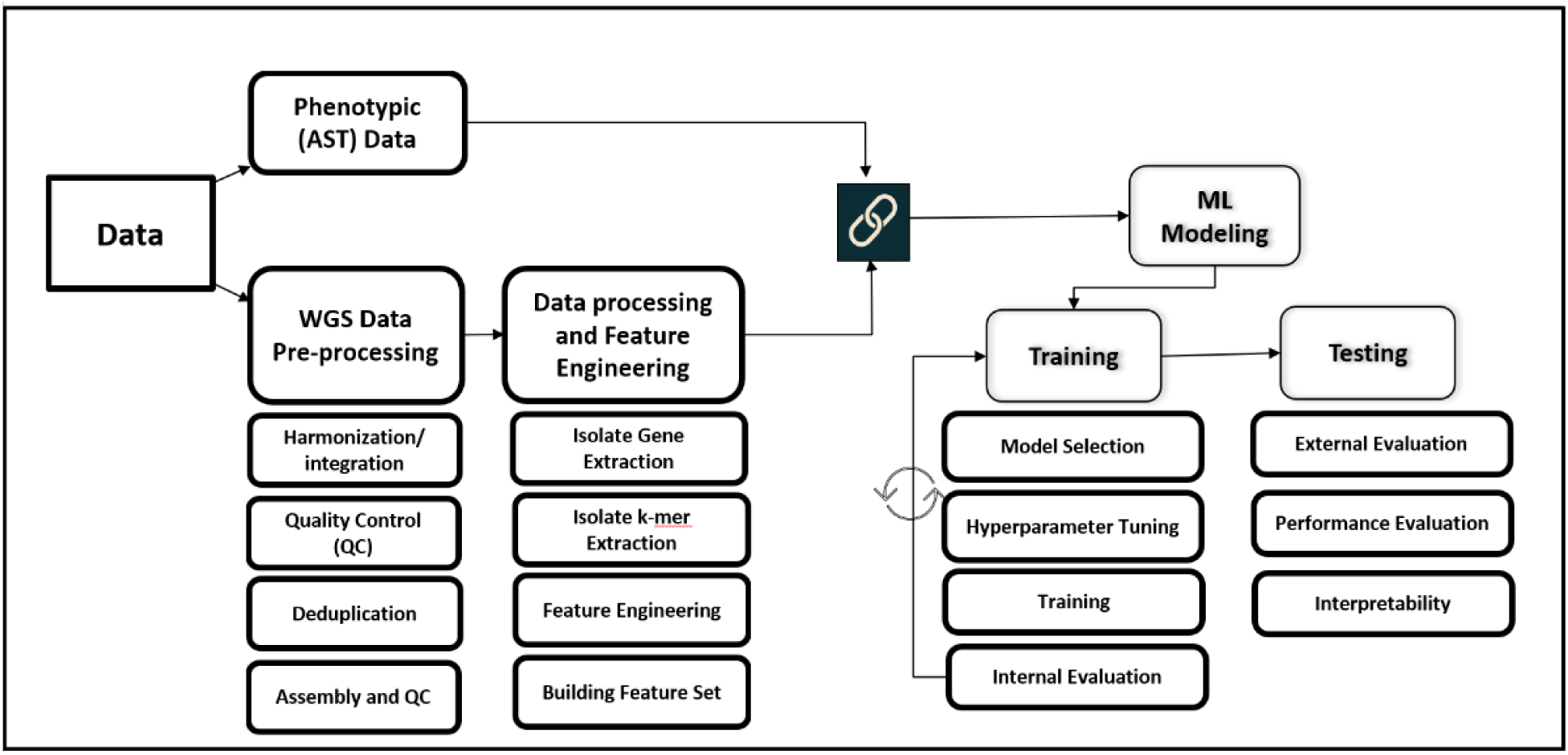
Workflow of data processing and machine learning modeling for ciprofloxacin resistance prediction in *Shigella*. Phenotypic AST data and WGS data were integrated following preprocessing steps. Resistance-related genes were extracted and represented using k-mer–based features to construct the feature matrix. Machine learning models were then trained using an internal training workflow. Final model performance was assessed through testing and interpretability analyses.

**Figure 2.**
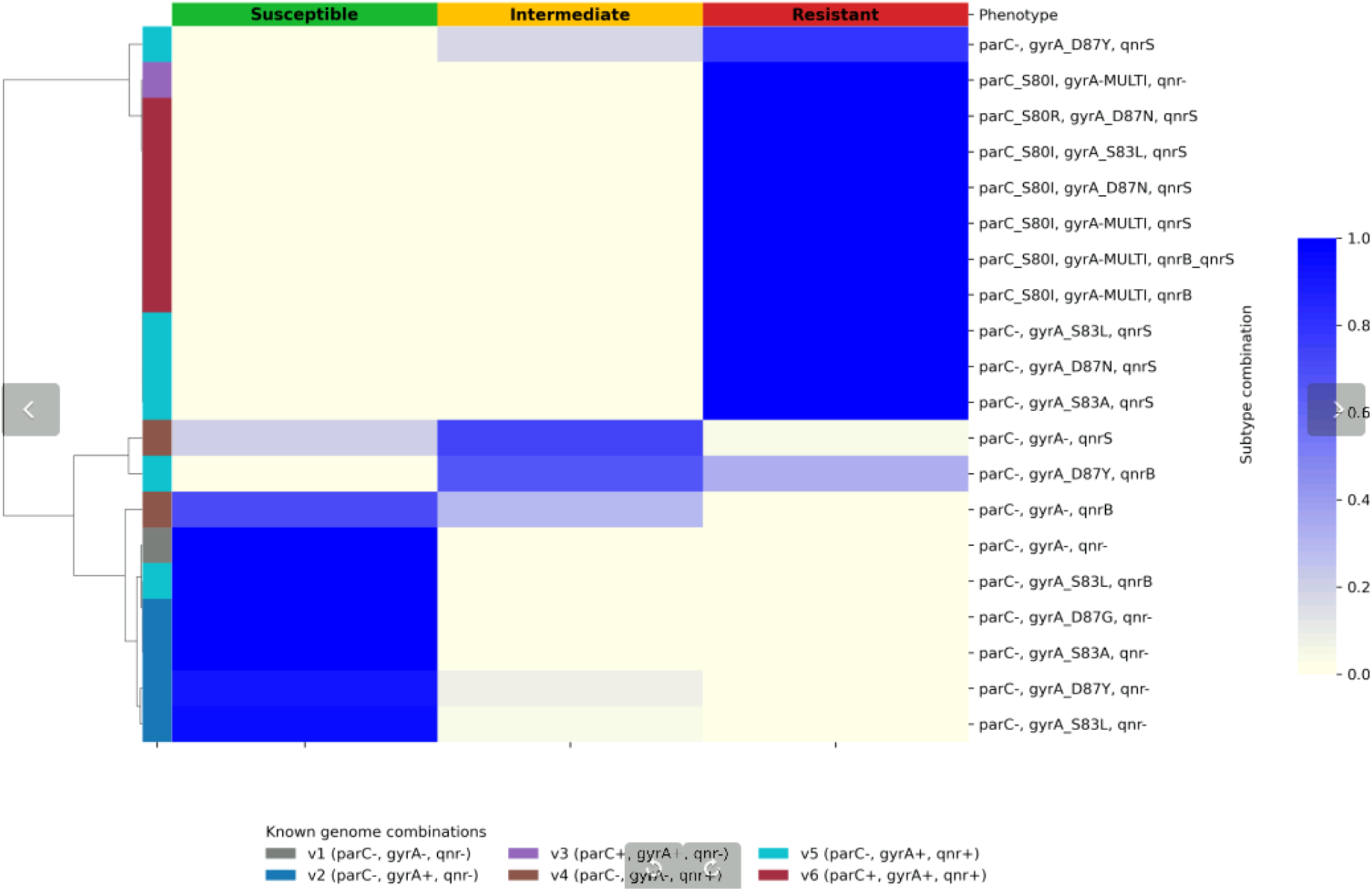
Distribution of QRDR mutation and plasmid-mediated resistance combinations across ciprofloxacin phenotypes in *Shigella*. Heatmap showing the number of isolates in association with the six ciprofloxacin phenotypes categorised based on combinations of known resistance determinants: ***par*C**, ***gyr*A**, and ***qnr***. These are v1 (*parC-, gyrA-, qnr-*), v2 (*parC-, gyrA+, qnr-*), v3 *(parC+, gyrA+, qnr-)*, v4 *(parC-, gyrA-, qnr+)*, v5 *(parC-, gyrA+, qnr+)*, v6 *(parC+, gyrA+, qnr+).* Each phenotype is further subdivided into 1, 4, 1, 2, 6, and 6 sub-phenotypes, respectively. Resistant isolates are strongly associated with canonical QRDR mutation combinations, particularly substitutions in *gyrA* (e.g., S83 and D87) often occurring alongside *parC* mutations and, in some cases, plasmid-mediated determinants. In contrast, susceptible isolates are predominantly characterized by wild-type or single-mutation profiles. Intermediate isolates display mixed patterns.

**Impact of k-mer Lengths on Model Performance:** Figure 3 compares the predictive performance of the RF model across different k-mer lengths, while performance results for the other models are summarised in the Supplementary Tables. Discrimination and calibration performance varied only modestly across k-mer lengths. AUC values remained consistently high for all k values (0.970–0.976), with k = 11 achieving the highest AUC. Similarly, Brier scores were low and comparable across k-mer lengths (0.035–0.037), with k = 11 yielding the lowest score, indicating slightly improved probability calibration. Model performance was largely consistent across k-mer lengths, suggesting that predictive accuracy was robust to the choice of k. The marginal improvement observed for k = 11 likely reflects efficient capture of mutation-associated sequence patterns within the targeted resistance genes. In contrast, threshold-dependent metrics, including F1 score, recall, precision, ME, and VME were identical across k-mer lengths, indicating that classification performance at the selected decision threshold was unaffected by k. Based on its marginally superior discrimination and calibration performance, k = 11 was selected as the optimal k-mer length for subsequent analyses.

**Figure 3.**
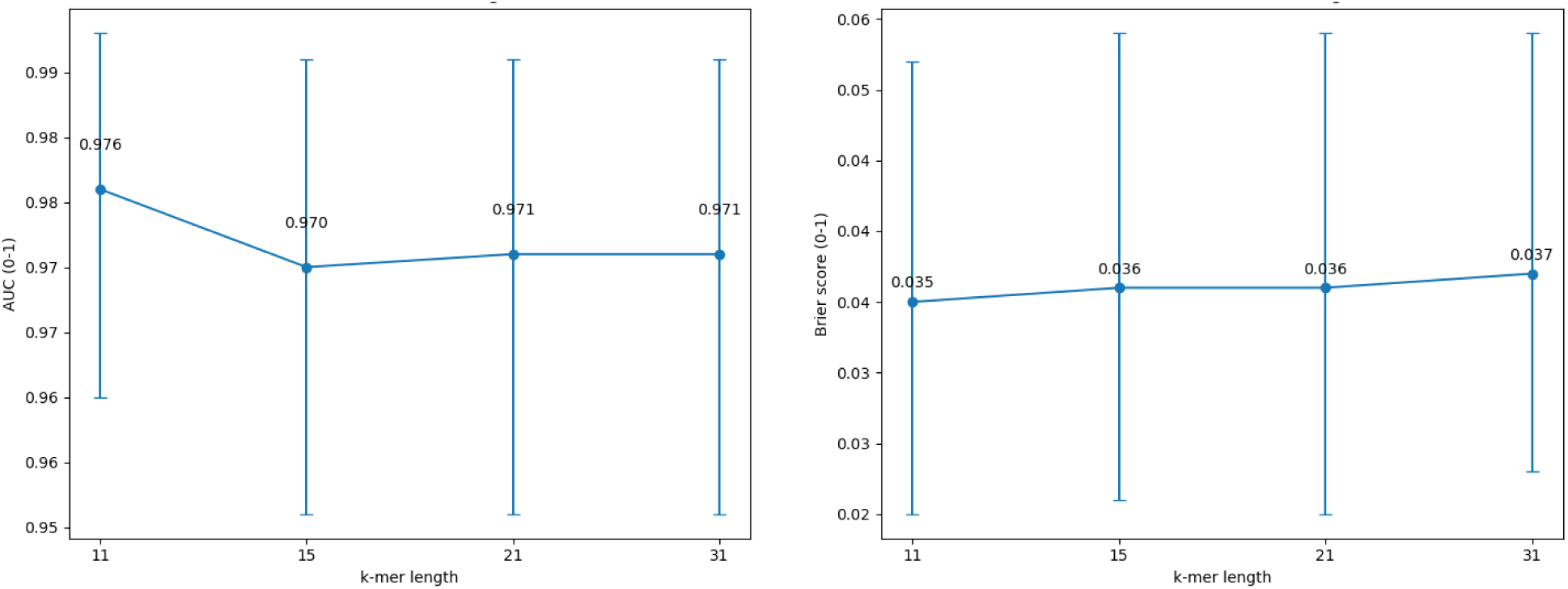
Performance comparison of Random Forest across k-mer lengths (k = 11, 15, 21, 31) for ciprofloxacin resistance prediction in *Shigella*. The left panel shows AUC (95% confidence intervals), and the right panel shows Brier score (95% confidence interval). Both metrics range from 0 to 1, Higher AUC values indicate improved discrimination, whereas lower Brier scores reflect better probabilistic calibration. Discrimination and calibration remained highly consistent across k values; however, k = 11 achieved the highest AUC and the lowest Brier score.

**Model Performance Evaluation:** Figure 4A presents the confusion matrices for all four models. For chromosomal models, RF and XGBoost produced identical predictions. SVM showed a slightly higher false positives (FP), whereas LR achieved the lowest FP. The inclusion of plasmid-mediated *qnr* determinants alongside chromosomal determinants (chromosomal-plasmid model) enhanced prediction performance across models. For RF and XGBoost, FP decreased from 3.8% (8/209) to 1.4% (3/209). False negatives (FN) decreased from 10.4% (15/144) to 7.6% (11/144), with corresponding increases in both true negatives (TN) and true positives (TP). SVM showed modest improvement, with the FP rate decreasing from 4.8% to 4.3% and the FN rate from 10.4% to 9.7%, whereas LR classification remained unchanged.

**Figure 4.**
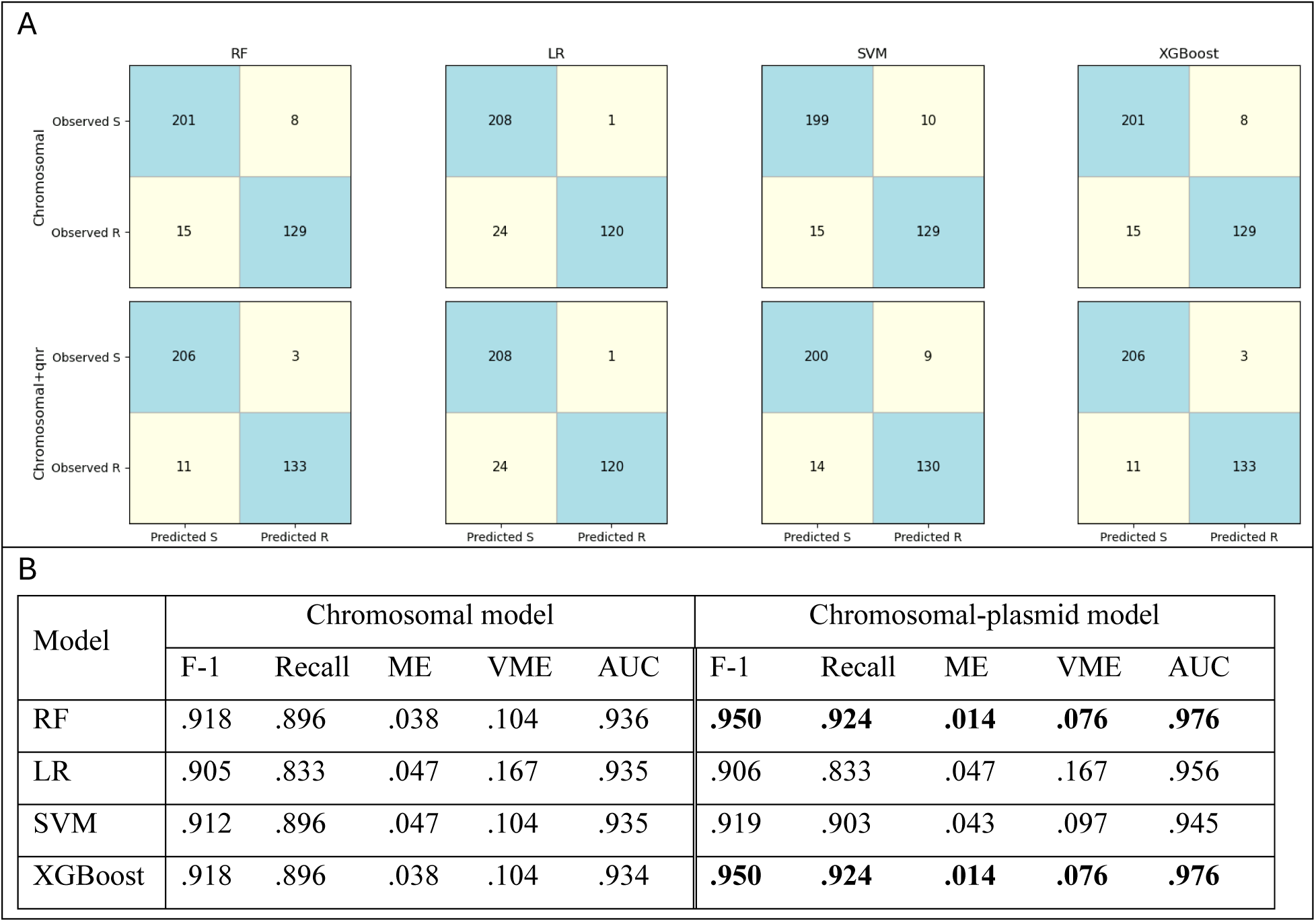
**Predictive performance of machine learning models for ciprofloxacin resistance in *Shigella* using k-mer features (k = 11)**. (A) Confusion matrices illustrating classification performance. Models in the top row are based on chromosomal determinants (*gyr*A, *par*C), whereas the bottom row incorporate both chromosomal and plasmid-mediated determinants (*qnr*). LR: logistic regression; RF: random forest; XGBOOST: extreme gradient boosting; SVM: support vector machine. Best metrics are highlighted. (B) Summary performance metrics. F-1 score (the harmonic mean of precision and recall), recall (the proportion of truly resistant isolates correctly identified), ME (major error rate, proportion of susceptible isolates incorrectly predicted as resistant), VME (very major error rate, proportion of resistant isolates incorrectly predicted as susceptible), and AUC (area under the curve, a measure of the model’s ability to distinguish between resistant and susceptible isolates across all classification thresholds), comparing chromosomal-only models with models incorporating plasmid-mediated markers.

Figure 4B presents key summary metrics. Chromosomal-plasmid models yielded higher F1 scores and recall reduced rates of ME and VME, and modest increases in AUC values compared with chromosomal-only models. RF and XGBoost achieved comparable performance across both models, whereas LR and SVM showed slightly lower performance for selected metrics.

**Feature Importance and Interpretability Assessment:** Given reduced ME and VME and comparable discrimination, the chromosomal–plasmid RF model was selected for SHAP-based interpretability analysis. SHAP was applied to the top-ranked k-mers contributing most strongly to model predictions (Figure 5). The summary plot illustrates the contribution of individual k-mers to model output across isolates. Each point represents the SHAP value of a given k-mer in a single isolate; with colour indicating feature values (red indicates presence of feature or higher impact on model; blue indicates absence or lower impact on model. The horizontal axis reflects the magnitude and direction of each k-mer’s contribution to predict resistance or susceptible and the y-axis represents the annotated genomic positions of the k-mers. Highest impact features were predominantly concentrated within well-characterized QRDR regions of *gyr*A and *par*C. In particular, k-mers spanning canonical resistance-associated amino acid substitutions (*gyr*A D87N and *gyr*A S83L and *par*C S80I) shoed the strongest contribution to model predictions. For example, k-mers mapping to *gyr*AD87N (AA position 87 and adjacent windows) exhibited strong positive SHAP values when present, indicating a strong association with resistance.

**Figure 5.**
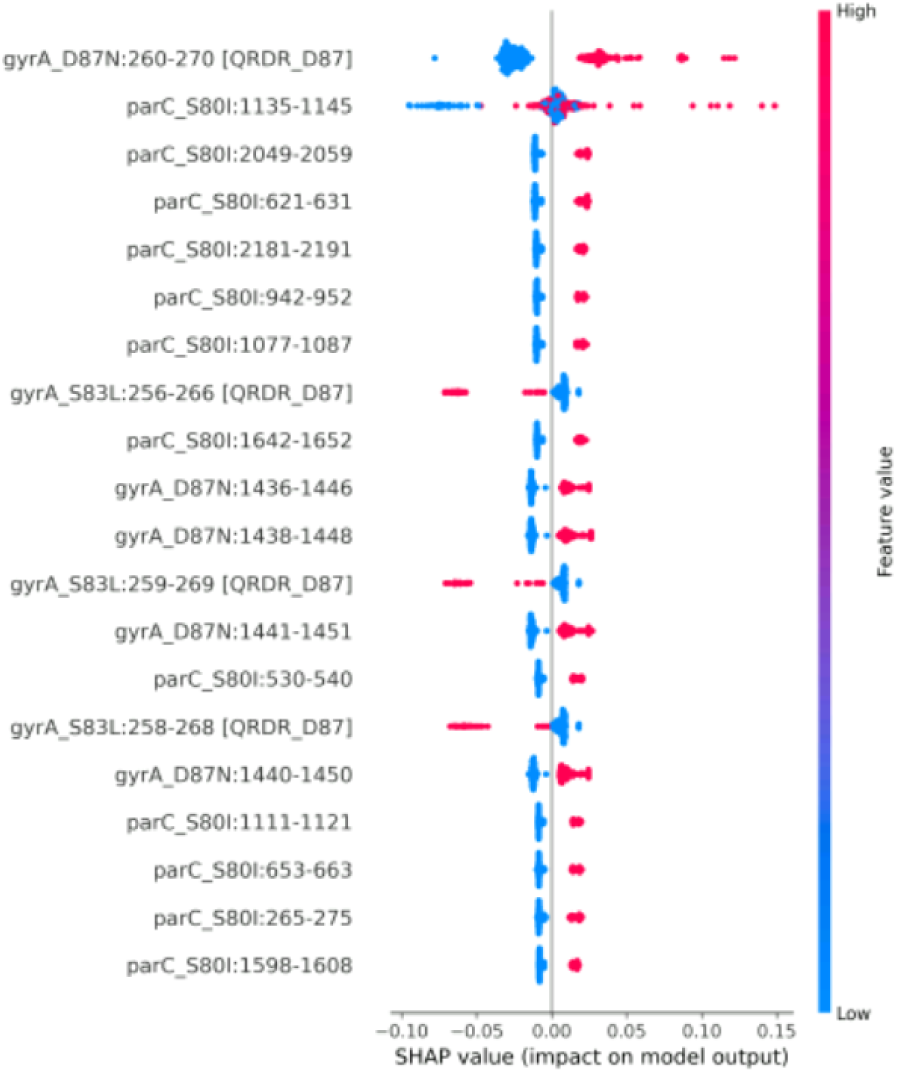
**SHAP summary plots illustrating k-mer feature contributions to ciprofloxacin resistance predictions in the chromosomal-plasmid model using 11-mers**. Each point represents a k-mer feature (length=11) feature for an individual isolate. Features are ordered by their overall importance (mean absolute SHAP value). Colour indicates feature value, with red indicates presence (higher feature value), and blue indicates absence or lower feature value. Positive SHAP values indicate features contributing toward resistance classification, whereas negative SHAP values indicate contributions toward susceptibility.

Similarly, overlapping k-mers covering *par*CS80I (e.g., nucleotide positions 621–631 and neighboring intervals) demonstrated directional effects toward resistance prediction. Notably, clusters of *parC*-derived k-mers with similar SHAP values were observed, appearing as groups of features with comparable importance, likely reflecting k-mers capturing the same underlying mutation signal within the QRDR region.

## Discussion

In this study, we developed an interpretable ML framework to predict ciprofloxacin resistance in *Shigella* using k-mer features derived from genomic resistant determinants, including chromosomal QRDR mutations in *gyrA*, and *parC* and plasmid-mediated *qnr* genes. We evaluated four supervised algorithms across multiple k-mer lengths and compared two feature sets: chromosomal determinant along and a combined chromosomal-plasmid model. RF algorithm demonstrated the most robust and consistent discrimination, and the inclusion of *qnr* alongside *gyrA* and *parC* improved predictive performance compared with chromosomal-only models. Based on these results, the chromosomal-plasmid RF model with k = 11 was selected as the optimal for predicting AMR phenotypes in *Shigella,* while retaining interpretability of known genomic determinants.

Importantly, model interpretability analysis confirmed that predictive performance was driven by well-characterized resistance mechanisms. High impact k-mers were concentrated within QRDR regions of *gyr*A and *par*C, particularly those spanning canonical substitutions such as S83L and D87N in *gyr*A and S80I in *par*C. These finding reinforce the central role of chromosomal mutations in ciprofloxacin resistance and demonstrate that k-mer based models can capture fine-scale sequence variation. The additional contribution of *qnr* suggests that plasmid-mediated mechanisms provide complementary, albeit more modest, predictive signal within this dataset, consistent with the role of *qnr* conferring low-level resistance that may facilitate or enhance the effects of QRDR mutations (18). We further evaluated the impact of k-mer length on predictive performance. Although performance differences across k values were modest, k = 11 achieved improved discrimination (highest AUC) and probability calibration (the Brier score) relative to longer k-mers (Figure 3). This result is consistent with previous studies suggesting that shorter k-mers provide an optimal balance between discriminatory resolution and redundancy (17, 20). For example, (17) selected k = 11 to retain informative sequence variation while limiting computational burden. In contrast, longer k-mers (k=21 or 31) may improve sequence specificity and genomic localization but can introduce sparsity, and increase feature dimensionality in practice by generating more unique and less shared sequence patterns across isolates, resulting in reduced feature overlap (21, 22).

A key strength of this study is the use of a targeted, biologically informed genomic feature space. By restricting analysis to k-mers derived from *gyrA*, *parC*, and *qnr*, we focused on the principal chromosomal and plasmid-mediated mechanisms of ciprofloxacin resistance in *Shigella*. This hypothesis-driven approach evaluated sequence-level variation within established resistance mechanisms and enhanced biological interpretability. This approach also reduced the risk of spurious associations that can arise in high dimensionality genomic feature spaces, where correlated features may capture lineage associated signals rather than causal resistance determinants (23). Previous studies have successfully inferred AMR phenotypes using known resistance genes, demonstrating high predictive accuracy across multiple pathogens (14, 24).

Although genome-wide k-mer models have shown strong predictive performance (13, 25), they may capture indirect or lineage-associated signals. In contrast, our targeted approach enables detection of subtle nucleotide-level variation within validated QRDR regions while maintaining biological interpretability and relevance.

Across the evaluated algorithms, predictive performance was broadly comparable, with RF and XGBoost achieving higher overall metrics than LR and SVM. This aligns with prior studies indicating that tree-based ensemble methods consistently demonstrate strong performance in genomic and clinical prediction tasks (26, 27). Given the comparable performance between RF and XGBoost, RF was selected due to its lower hyperparameter complexity, greater stability (28), and more readily interpretable feature importance measures (29), facilitating biological interpretation of resistance-associated k-mers (30, 31). Although ML approaches show promise for AMR prediction, several methodological considerations warrant attention. First, ML models may struggle to learn robust and generalizable patterns when key biological information is incomplete or when resistance arises from complex cellular and evolutionary mechanisms (26). Second, AMR is closely linked to clonal population structure. AMR is often driven by the global expansion of multidrug-resistant lineages (32), resulting in a close association between clade membership and resistance phenotype. Because resistant clades vary across time and geographic regions (32), model performance can be heavily influenced by local epidemiology and may not generalize across geographic regions or time periods (33). Third, many ML approaches treat genetic features independently, whereas resistance phenotypes often result from non-linear interactions among genetic and environmental pressures, including antibiotic exposure and ecological selective forces that shape resistance evolution (18, 34). Although methods exist to model such interactions, the combinatorial complexity of genomic features presents substantial analytical challenges (26).

Overall, our study demonstrates several strengths. It leverages a comprehensive and integrated provincial dataset of *Shigella* isolates collected through routine public health surveillance in Ontario, providing representative coverage of circulating strains and resistance patterns. The use of whole-genome sequencing enabled detailed characterization of both chromosomal and plasmid-mediated resistance determinants, supporting robust evaluation of genomic predictors of AMR. In addition, this work contributes to the growing body of evidence supporting the application of interpretable ML approaches for genomic AMR prediction.

The findings should be interpreted in light of several limitations. The analysis was restricted to k-mers derived from a limited set of known resistance determinants (*gyrA*, *parC*, and *qnr*), and additional mechanisms such as efflux pump regulation, promoter variation, or novel resistance loci may also contribute to ciprofloxacin resistance (19). The reliance on k-mers alone may not capture all biologically relevant variation; integration of complementary feature types, such as SNPs or gene presence or absence metrics could improve predictive performance and biological resolution. Previous studies have shown that integrating multiple feature types can enhance predictive performance and biological interpretability. For example, Liu, Wang (17) demonstrated that integrating SNP and k-mer features improved ML model performance for predicting ciprofloxacin resistance in *Staphylococcus aureus*, with the integrated model achieving a higher AUC (0.986) than models based on SNPs alone (0.984) or k-mers alone (0.955). Although tree-based models such as RF can capture non-linear relationships between features, this study did not explicitly model epistatic interactions between resistance determinants. Interactions between chromosomal mutations and plasmid-mediated determinants may influence resistance phenotypes beyond additive effects (35, 36). Finally, as models trained on isolates from a single geographic region, Ontario, Canada, population structure and local epidemiology may limit generalizability to broader *Shigella* populations (23).

In conclusion, this study demonstrates that integrating biologically informed k-mer features with interpretable ML approaches can effectively capture key genetic mechanisms underlying ciprofloxacin resistance in *Shigella*. The results highlight the predominant role of chromosomal QRDR mutations while clarifying the complementary contribution of plasmid-mediated markers. Together, these findings support the value of combining genomic resolution with transparent modelling to advance resistance prediction and mechanistic understanding in public health surveillance.

## Material and Methods

### Ethics Statement

This project has received ethics review clearance from Public Health Ontario (PHO)’s Ethics Review Board (ethics file number: 2025-005-01). The need for informed consent was waived by Public Health Ontario’s Ethics Review Board as per Article 5.5B of the Tri-Council Policy Statement: Ethical Conduct for Research Involving Humans (TCPS2), Canada’s national research ethics policy, which states that researchers are not required to seek participant consent for research that relies exclusively on the secondary use of non-identifiable information.

### Data sources and dataset preparation

We analyzed de-identified whole genome sequences (WGS) and antimicrobial susceptibility testing (AST) data from *Shigella* spp. isolates recovered at PHO, the provincial reference laboratory in Ontario, Canada between 2018 and 2025. Clinical specimens are submitted to PHO by community and hospital laboratories across the province, where *Shigella* isolates are recovered and characterized through identification, serotyping, and AST for standard antimicrobial agents, and sequenced at the National Microbiology Laboratory (NML), as part of routine public health surveillance.

### Phenotypic AST data

AST data for *Shigella* isolates were extracted from the PHO Laboratory Information Management System (LIMS). PHO routinely performs phenotypic testing on isolates submitted through provincial surveillance, encompassing the majority of confirmed shigellosis cases in Ontario. Testing was conducted in accordance with current Clinical and Laboratory Standards Institute (CLSI) guidelines, and resistance classifications were determined using minimum inhibitory concentration (MIC) breakpoints defined in CLSI M100-ED33:2024 (37). Phenotypic AST results were available for antimicrobial agents commonly used to treat shigellosis, including ampicillin, azithromycin, ceftriaxone, ciprofloxacin, and trimethoprim-sulfamethoxazole(5). However, th**e** present study focused on ciprofloxacin because its resistance in *Shigella* represents a growing global concern and is a key indicator reported in the WHO Global Antimicrobial Resistance Surveillance System (GLASS)(38).

### Whole Genome Sequencing (WGS) data

WGS of enteric bacterial isolates was performed at the NML. RNA-free genomic DNA was extracted from cultured isolates and prepared for sequencing using Nextera library preparation, generating adapter-ligated DNA fragments (300 bp). Libraries were sequenced on Illumina MiSeq or NextSeq platforms, where DNA fragments were cluster-amplified on a flow cell, imaged, and processed through base calling, filtering, and quality scoring. Post-run data underwent quality control assessment, including read metrics and coverage evaluation prior to downstream analysis. Sequence data were analyzed using validated bioinformatics pipelines within the Integrated Rapid Infectious Disease Analysis (IRIDA) and BioNumerics platforms, including whole genome multi-locus sequence typing (wgMLST) for cluster analysis and relatedness assessment, and organism-specific pipelines such as the Salmonella In Silico Typing Resource (SISTR) and ECTyper for *in silico* serotype prediction.

WGS was performed on de-identified specimens as part of routine public health surveillance activities.

### K-mer Feature Selection

Ciprofloxacin resistance is primarily mediated by point mutations in the chromosomal genes *gyrA* and *parC,* as well as plasmid-borne determinants such as *qnr* (19). Accordingly, k-mer feature selection derived from these established resistance loci (*gyrA*, *parC*, and *qnr*).

### Data Processing Pipeline

A custom bioinformatics pipeline was implemented to ensure data quality and extract AMR-related features (Figure 1). Raw paired-end reads were quality filtered using *fastp* (v0.23.2) to remove adapter sequences, trim low-quality bases (Phred score <20), and discard short reads (<50 bp). To reduce biases from over-represented sequences, deduplication was performed using BioNumPy. Filtered reads were assembled into contigs using *Shovill* (v1.1.0) with default parameters. Assemblies failing to meet minimum quality thresholds (N50 ≥ 50 kb or total assembly length > 4 Mb) were excluded.

The resulting assemblies were screened with AMRFinderPlus (v4.0.23; NCBI AMR database) to identify known AMR genes and resistance-associated point mutations. Gene presence/absence calls were compiled into a binary gene-by-sample matrix. This matrix was merged with phenotypic ciprofloxacin susceptibility data. After quality control, 1,424 isolates met inclusion criteria for analysis, including 577 (40.5%) resistant and 660 (46.4%) susceptible isolates.

Intermediate isolates (n = 187, 13.1%) were grouped with the susceptible category to enable binary classification, given the relatively small number of intermediate cases and variability in the interpretation of this category (39).

### K-mers Feature Extraction

To extract sequence features associated with ciprofloxacin resistance, BioSQI was used to generate isolate-level k-mer profiles for the target genes *gyr*A, *par*C, and *qnr*. K-mers were generated using a sliding window approach and encoded as binary presence-absence features. To assess the effect of sequence resolution on model performance, multiple k-mer lengths (k = 11, 15, 21, and 31) were examined.

K-mers were generated separately for each *Shigella* isolate and subsequently aggregated into a unified k-mer feature matrix, where rows correspond to isolates and columns represent distinct k-mers. Let 𝑀∈^𝑛×𝑝^ denote the resulting binary matrix, where 𝑛 represents the number of isolates and 𝑝 represents the total number of unique k-mers. Each element 𝑀_𝑖𝑗_ indicates the presence of k-mer 𝑘_𝑗_ is in isolate 𝑖, such that:

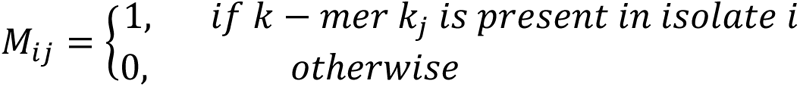

### Feature Selection

Given the high dimensionality of the k-mer presence–absence matrix, which can lead to computational inefficiency and overfitting (26), feature selection was performed using LASSO (Least Absolute Shrinkage and Selection Operator) regularization (40).

Specifically, a logistic regression model with an ℓ_1_- penalty was fitted to the binary ciprofloxacin resistance outcome, minimizing the objective function:

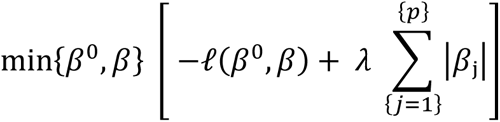

where ℓ(⋅) denotes the log-likelihood function, 𝛽 represents the regression coefficients associated with the k-𝜆 features, and ℓ_1_is the regularization parametercontrolling the degree of sparsity. The ℓ-penalty shrinks some coefficients exactly to zero, thereby selecting a subset of informative k-mers. The resulting reduced k-mer feature set was used for downstream ML training.

### ML model Construction

Supervised ML models that are commonly used in genomic prediction were applied, including Logistic Regression (LR), Random Forest (RF), Support Vector Machine (SVM), and Extreme Gradient Boosting (XGBoost). Two gene-based model sets were constructed. **Model Set 1 (chromosomal model)** included models trained using k-mers derived exclusively from the chromosomal loci *gyr*A and *par*C. **Model Set 2 (chromosomal-plasmid model)** included models trained using k-mers derived from *gyr*A, *par*C, and the plasmid-mediated resistance gene *qnr*. RF, SVM, and LR models were implemented using the Scikit-learn Python package, with hyperparameters optimized via grid search within 5-fold cross-validation on the training dataset. The XGBoost model was implemented using the XGBoost library, with hyperparameters tuned using cross-validation. The dataset was randomly divided into training (75%) and testing (25%) sets. Model performance was evaluated on the independent testing dataset. Following model development, the most predictive k-mers identified across models were mapped back to their corresponding reference AMR genes to facilitate biological interpretation of the model outputs.

### Model Evaluation

Model performance was assessed by comparing predicted AMR genotypes from the testing set with corresponding AST results using multiple metrics, including precision (the proportion of predicted resistant isolates that are truly resistant), recall (the proportion of truly resistant isolates correctly identified), F1 score (the harmonic mean of precision and recall), and area under the receiver operating characteristic curve (AUC; a measure of the model’s ability to distinguish between resistant and susceptible isolates across all classification thresholds). In addition, AMR-specific error metrics were calculated, including major error (ME), and very major error (VME) (26). An ME occurs when a susceptible isolate is incorrectly predicted as resistant, whereas a VME occurs when a resistant isolate is incorrectly predicted as susceptible (37). Model calibration was evaluated using the Brier score (41), defined as the mean squared difference between predicted probabilities and observed binary outcomes (resistant = 1, susceptible = 0). The Brier score ranges from 0 to 1, with lower values indicating better agreement between predicted probabilities and observed outcomes. The Brier score was assessed alongside AUC to evaluate both the discriminative ability and calibration performance of models.

### Model Interpretation

To interpret model predictions and quantify the contribution of individual k-mer features to ciprofloxacin resistance classification, SHapley Additive exPlanations (SHAP) analysis was applied (29). SHAP values were computed to estimate the marginal contribution of each feature to the model prediction for individual isolates, enabling both global and local interpretability. Global feature importance was assessed by summarizing the absolute SHAP values across isolates, whereas isolate-level SHAP profiles were examined to determine how specific k-mers contributed to resistance or susceptibility predictions.

## Data and Code Availability

The Fastq files of *Shigella* spp. from Ontario, Canada included in this study have been deposited in the National Centre Biotechnology Information (NCBI) GenBank database. [Approval for data sharing is currently in progress, and accession numbers will be provided upon completion.]

## Data Availability

Approval for data sharing is currently in progress, and accession numbers will be provided upon completion.

## Acknowledgements

The authors gratefully acknowledge the staff and scientists at Public Health Ontario (PHO) and National Microbiology Laboratory (NML) for their support. We especially thank Analyn Peralta, Ashleigh Sullivan, Karthikeyan Sivaraman, Mark Horsman, and George S. Long for their assistance with laboratory and technical aspects of the study. MRG was supported by the CIHR Health System Impact Fellowship (HSIF). The funders had no role in study design, data collection and analysis, decision to publish, or preparation of the manuscript.

## Author Contributions

Conceptualization: MRG, SNP, JPH, VRD; Data curation: MRG, PZ, AV; Formal Analysis and Interpretation: MRG, LCR, SNP, JPH, VRD; Methodology: MRG, SNP, JPH, VRD; Visualization: MRG; Writing – original draft: MRG; Writing – review & editing: PZ, AV, LCR, SNP, JPH, VRD.

## Supplemental Material

**Supplementary Figure 1.**
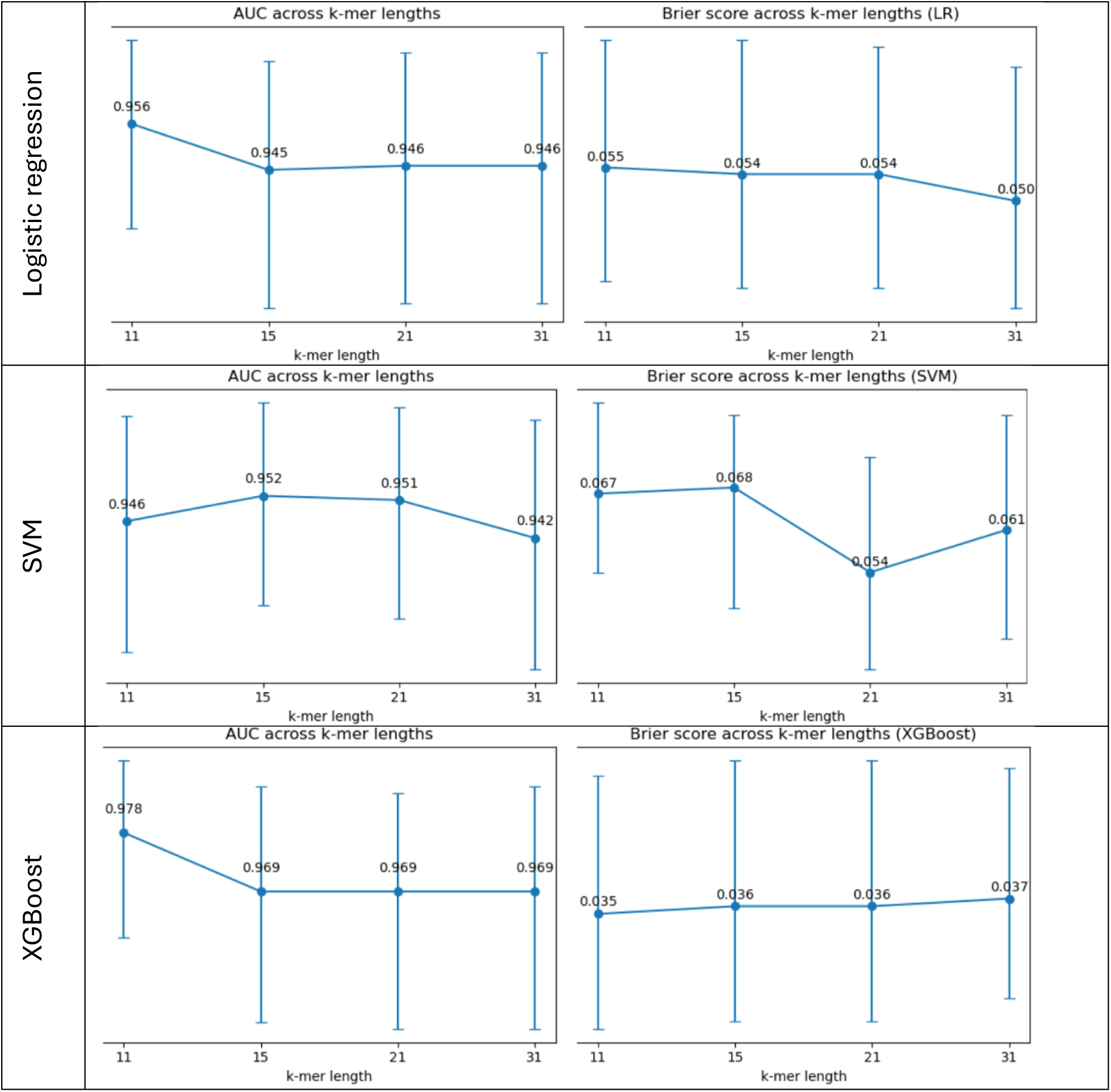
Performance comparison of LR, SVM, and XGBoost across k-mer lengths (k = 11, 15, 21, 31) for ciprofloxacin resistance prediction in *Shigella*. The left panels display AUC (95% confidence intervals), and the right panels display Brier score (95% confidence intervals). Consistent with the findings observed for the Random Forest model, discrimination and calibration remained fairly stable across k values, with k = 11 achieving the overall highest AUC for logistic and XGBoost and the lowest Brier score for XGBoost.

